# Understanding the impact of the COVID-19 pandemic on well-being and virtual care for people living with dementia and care partners living in the community

**DOI:** 10.1101/2020.06.04.20122192

**Authors:** P Roach, A Zwiers, E Cox, K Fischer, A Charlton, CB Josephson, SB Patten, D Seitz, Z Ismail, EE Smith, on behalf of the PROMPT Collaborators

**Affiliations:** Department of Community Health Sciences, University of Calgary; Hotchkiss Brain Institute, University of Calgary; O’Brien Institute of Public Health, University of Calgary; Department of Clinical Neurosciences, University of Calgary; entre for Health Informatics, University of Calgary; Cuthbertson & Fischer Chair in Pediatric Mental Health, University of Calgary; Department of Psychiatry, University of Calgary

## Abstract

The COVID-19 pandemic has necessitated public health measures that have impacted the provision of care for people living with dementia and their families. Additionally, the isolation that results from social distancing may be harming well-being for families, as formal and informal supports become less accessible. For those with living with dementia and experiencing agitation, social distancing may be even harder to maintain, or social distancing could potentially aggravate dementia-related neuropsychiatric symptoms. To understand the lived experience of social and physical distancing during the COVID-19 pandemic in Canada we remotely interviewed 21 participants who normally attend a dementia specialty clinic in Calgary, Alberta, during a period where essential businesses were closed and healthcare had abruptly transitioned to telemedicine. The impacts of the public health measures in response to the pandemic emerged in three main categories of experience: 1) personal; 2) health services; and 3) health status (of both person living with dementia and care partner). This in-depth understanding of the needs and experiences of the pandemic for people living with dementia suggests that innovative means are urgently needed to facilitate provision of remote medicine and also social interaction and integration.

## Introduction

With the emergence of COVID-19 (SARS-CoV-2) as a novel coronavirus in late 2019, a global pandemic has led to varying levels of social distancing restrictions across the world. This includes restrictions on public gatherings, the conduct of businesses, the accessibility of healthcare, and movement outside the home.

The unintended consequences of these measures may included disrupted routines for people living with dementia, increased stress on care partners as they manage the demands of caring for someone living with dementia, and difficulty satisfying essential needs such as obtaining groceries and medications without the typical social and health service support, or support from personal networks. These measures have the potential to cause social isolation, which is increasingly recognized to be detrimental to overall well-being (Vernooij-Dassen & Jeon, 2016; Ward et al., 2018) Well-being requires deep and meaningful human connections, health status above an expected baseline, positive interpersonal expectations, and a sense of self (Feeney & Collins, 2015). Maintaining well-being is a key component of person-centered dementia care (Kitwood, 1997). Moreover, the rapid move to virtual care provision may influence how and when people access treatment from healthcare providers, in addition to how providers can continue to provide high quality care in new virtual settings.

Multiple professional organizations including Alzheimer’s Disease International (2020) have issued emergent COVID-related advice for persons with living with dementia, but so far these recommendations have been based on little evidence, rarely collected systematically.To begin building an evidence base for the impact of the COVID-19 pandemic on well-being and healthcare of persons living with dementia, we conducted a qualitative study of persons living with dementia and care partners receiving community-based care at a dementia specialty clinic. A qualitative approach was used to collect rich data on lived experience that incorporated a reflexive thematic approach to content analysis.

## Research Design

We completed remote, in depth interviews using telephone with family members or care partners, as well as the person with dementia if they wished to and were able to provide consent. In order for participants to provide detail on the changes and impacts of the pandemic on their lives it is important that these data ae collected while the pandemic is occurring and while social distancing measures are in place.

### Recruitment

We purposely recruited participants (people living with dementia and family members/care partners) who had attended the Cognitive Neurosciences Clinic in Calgary, Alberta, Canada since January 1, 2019; had a diagnosis of dementia verified by a specialty neurologist or psychiatrist working in the clinic; were a participant in the PROspective Registry of Persons with Memory SyMPToms (PROMPT); and had previously provided written consent to be contacted about additional research. Snowball sampling was also used, with two dyads recruited this way, likely through a community network for younger people living with dementia. The Calgary Cognitive Neurosciences Clinic operates out of two large tertiary-care sites in Calgary, Alberta, Canada. It is the only dementia speciality clinic in Southern Alberta, with a catchment area of approximately 1.5 million. Patients are referred by a primary care provider and assessed for cognitive impairment and dementia by a staff of nurses and either a cognitive neurologist or psychiatrist. In Alberta, there is universal government funding for physician visits and diagnostic testing. PROMPT was established in the clinic in 2010 to investigate patient outcomes. Additional details on PROMPT design have been published previously (Sheikh et al., 2018). All patients seen at the clinic are eligible to participate and enrol in PROMPT. Participation includes the opportunity to provide consent to be contacted for future research.

Consent was obtained using Qualtrics survey software (Qualtrics, 2019) or explicit oral consent. Due to the nature of some of the clinic populations, explicit oral consent was necessary so that participants who wanted to participate but perhaps did not have access to the internet, email, or who had disablements preventing them from using devices or computer screens could still be included. Explicit oral consent provided a way to ethically include a diverse population. Process consent (Dewing, 2007) was used throughout the project, where all participants provided verbal assent at every research encounter (including the initial call to introduce the project and any subsequent contact including interviews) to ensure that the participants were comfortable participating at that day or time. Options of telephone or Zoom interviews were used as a technological tool to complete the interviews in order to adhere to social distancing mandates put in place in the province of Alberta when the public state of emergency was declared on March 17, 2020. Potential participants were then contacted by telephone and informed of the project, after which information sheets and consent forms were emailed or mailed to interested participants who had a minimum of 24 hours to consider the information and discuss it with family members or health providers before providing consent to participate and the interview is scheduled. This study was approved by the Conjoint Human Research Ethics Board at the University of Calgary (REB20–0559).

### Data Collection

In depth interviews were used as they are an appropriate way to gain a robust understanding of lived experience and allow for exploration of topics that are important to the participant, maintaining a participatory approach to working with people living with dementia and care partners. An interview guide was created asking general questions about experiences of isolation due to the pandemic and virtual health care experiences, and was tested and updated after the first two interviews to act as pilots. It was anticipated that theoretical saturation of participant experiences would be achieved after completion of 15–20 interviews and so a target of recruiting 20 participants was initially determined. In depth interviews were audio-recorded and field notes were taken during the interviews in a dedicated field journal, along with reflexive field notes written directly after the interviews. Demographic data was also recorded and was available through the current registry, and information on whether the participants had contracted Covid-19 or were tested for SARS-CoV 2 was collected by interview. To collect the planned sample size of 20 completed interviews, we contacted 45 people of whom 21 consented and completed the interview. Interviews ranged in duration from 8m:33s to 34m:38s (mean length = 21m:12s) and all interviews were completed over the phone as per participant request. Interviews were conducted between April 23 and May 21, 2020, with recruitment and data collection completed by KF, AZ and EC, all research staff with extensive experience working with people living with dementia. The interviewers were unknown to the participants at the time of interview but introduced themselves and the reason for the work at initial contact and at the time of interview.

In Canada, provincial governments are primarily responsible for implementing public health measures for pandemics. A timeline of provincial measures for social distancing and health system reorganization was created by review of public records of the City of Calgary and government of Alberta, communications from Alberta Health Services (the single public healthcare system in Alberta that manages the Cognitive Neurosciences Clinic), and the Calgary Herald newspaper (Calgary, Canada). From March 4 onwards, the provincial Chief Medical Officer of Health conducted daily briefings on virus activity and the government response. The Cognitive Neurosciences Clinic ceased routine in-person visits on March 13 and virtual care appointments for the regular attendees of the Cognitive Neurosciences Clinic were implemented on March 14, 2020. A timeline of public health measures implemented by the government of Alberta is shown in Figure 1.

**Figure 1:**
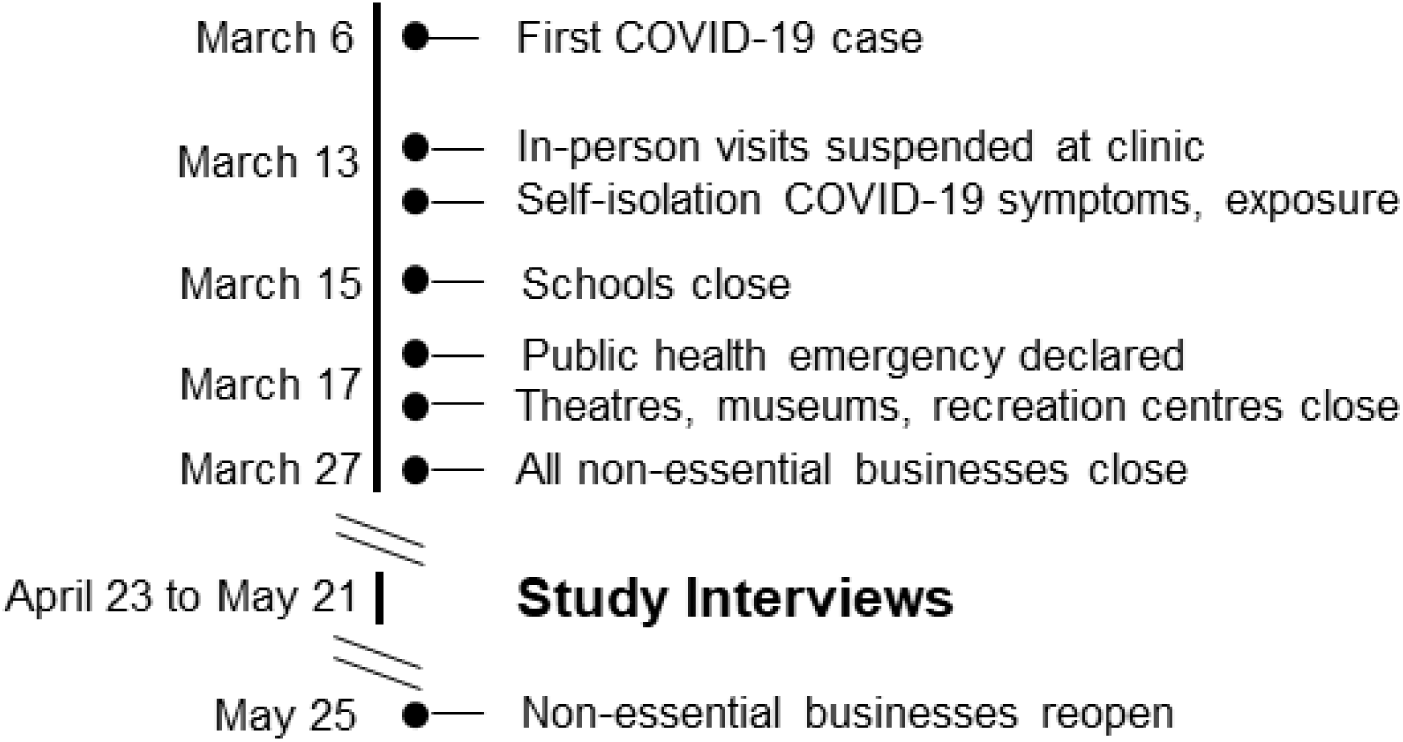
Timeline of COVID-19 Public Health Response in Alberta, Canada.

### Analysis

Audio recordings were transcribed verbatim using a secure professional transcribing service, and anonymized and verified by the research team. Transcripts and field notes were managed using NVivo 12 (QSR International, 2018) for the qualitative analysis and data management. Interviews were transcribed concurrently while data collection was still occurring so that generated themes could be discussed at subsequent interviews to achieve theoretical saturation. Immersion in the qualitative data was the first step of data analysis. Transcripts were read and re-read while the interview recordings are replayed. A reflexive thematic analysis (Braun et al., 2016; Clarke et al., 2019) was used to analyse the interview and field note data. Reflexive thematic analysis positions the researcher as a mechanism in the production of the analysis. This form of analysis also requires a cognizance of theoretical assumptions that centers the experience of the person living with dementia and care partner, and their experiences within the data and the analytical process. Analytical thoughts and iterative analyses were captured using memos and annotations. Disagreements over analyses and emergent themes were resolved via team discussion. Themes and subthemes were brought back to the participants for member checking and to enhance qualitative rigor and establish trustworthiness of the data (Lincoln & Guba, 1985).

## Results

### Participant Demographics

There were 20 dyads of persons living with dementia and care partners; in 18 cases the persons living with dementia declined to participate but the care partner participated and in one case the person living with dementia and care partner each participated on their own. Diagnoses and last Folstein Mini Mental Status Examination (Folstein et al., 1975) scores for the persons with dementia are shown in Table 1. No participants reported a diagnosis of COVID-19 for anyone living in their household; two care partner participants suspected they may have had COVID-19 due to symptoms but were not tested; and a third care partner participant reported being told to isolate due to being in a high risk group of complication from COVID-19.

**Table 1.**
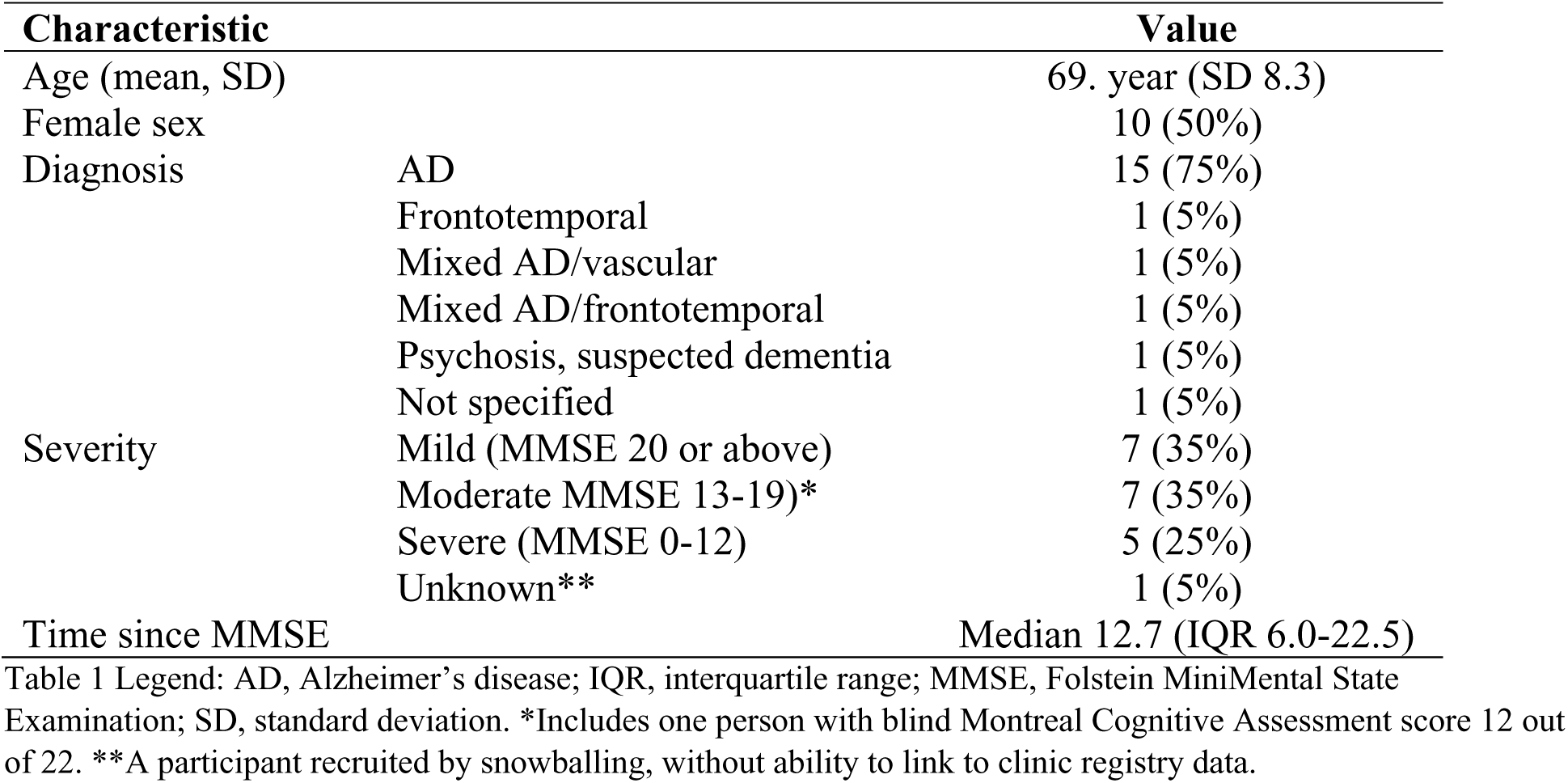
Characteristics of the twenty persons living with dementia who were interviewed or whose care partner was interviewed.

### Thematic Analysis: Experiences of the COVID-19 Pandemic

Through the completion of the qualitative analysis with coding and synthesis completed by AZ, EC and directed by PR (Principal Investigator and qualitative methods expert), three overarching themes emerged from the data (see Table 2) that clearly demonstrated the impact of the COVID-19 pandemic on experiences of cognitive neuroscience patients and their care partners. The themes built on a coding tree that started with small, descriptive codes about discrete areas of day to day life and were then synthesized to larger descriptions of experience. These themes are described in more detail below and have edited only for general grammar and spelling to enhance readability.

**Table 2:** Experiences of the COVID-19 Pandemic.

| Theme | Subthemes |
| --- | --- |
| **Personal Impact of Public Health Measures** | Personal Support |
|  | Managing day to day activities |
| **Health Services Impact of Public Health Measures** | Home Care Services & Support |
|  | Community Programming |
|  | Virtual Medicine: barriers and facilitators |
| **Health Status Impact of Public Health Measures** | Anxiety and Mental Health Concerns |
|  | Social distancing and cognitive decline |
|  | Care partner burnout |

### Personal Impact of Public Health Measures

#### Personal Support

Participants noted significant changes in their experiences of social support during the pandemic. Social and physical distancing, as enacted by public health measures, meant that the normal informal family and community supports were limited.

> *“So basically, being confined to the home, in general. Obviously, at the start the weather also wasn’t great, so, basically, the combination of, you know, bad weather, being… completely confined to the house, and then basically, the social isolation for my mom personally, that has been the biggest toll for her.” (Participant 124; Care Partner)*
>
> *“There’s, there’s no method to cope. I can’t ask people, family to come and help me. I can’t, there has been nothing I can do. No, I have not found anything for me personally that has been good about it. I have not been able to go to work. I have no socialization. I’m just left to cope on my own.” (Participant 141; Care Partner)*

It was also clear that the informal support provided by the community or by family members was crucial for many people who were anxious or afraid to leave the house for essentials. Those participants who had family support reinforced these messages.

> *“The first time we did something at the superstore, we put in a pick-up order. I don’t have a cell phone. I have a computer but not a cell phone. So my daughter said ‘You know, you put your order in and then I’ll go over and pick it up when it’s ready.’ She would do that for me. And we social distanced the whole time, like she’d drive up in the driveway.” (Participant 123; Care Partner)*

#### Managing day to day activities

Many care partners reported stress around coping with day to day activities of living and some of this stress was due to no longer having the formal home care service they had been receiving before the pandemic and shut down of services.

> “*That’s when I got him to take a bath as well. But it was a long time. I still have to coax him. He’s been changing… like, I’ve ironed clothes for him the other day… But he’ll just put on whatever and then he’ll wear it for days…” (Participant 115; Care Partner)*
>
> *“I don’t know that we are coping that well. (laughs) Um, there really was nothing I can do. I’m just trapped at home with him. I’m trapped at home.” (Participant 141; Care Partner)*
>
> *“I think probably one of the biggest challenges for my husband because of his cognition is the social distancing part. We had friends that sat about ten feet away and we were on the other side of our deck. And so when they came he was automatically wanting to go up and greet them and shake his hand… So I think sometimes it’s, the social distancing … Um, I don’t know. I think with the level of cognition that, uh, some days my husband has said it’s a foreign concept – like social distancing… I would not say that he understands that.” (Participant 106; Care Partner)*

### Health Services Impact of Public Health Measures

#### Health Care Services & Support

The personal impact of the pandemic and associated public health measures is closely associated to the health services impact of these same measures. Participants reported feeling alone and unsupported in many cases and were largely understanding of the need for health services to focus on the pandemic response. It was also clear, however, that some degree of ongoing contact was crucial for continuing to support families living with dementia.

> *“That’s kind of what I feel is, ‘You’re just on your own.’…I haven’t heard from anyone. I finally phoned someone the other day and ‘Yeah. Well people in AHS [Alberta Health Services] have been redeployed’, etc. etc., which is just fine. So maybe it would have been helpful if like your caseworker, somebody just to check in. Yeah, for sure.” (Participant 105; Care Partner)*
>
> *“Well, there’s nothing much because right now home care isn’t coming into the home. So the one on one, you know, he’s not getting that. If that was still happening I think that would be a huge help. But, it’s not and I can’t, you know, I totally understand and I don’t begrudge it. I want everybody to be safe but it is a very isolating, difficult time.” (Participant 103; Care Partner)*

Some participants did report formal service and community programming continuing to stay in contact and support families in innovative ways, and this continued contact was appreciated, even if the service level and model delivery had changed when the program had to stop in-person programming.

> *“He goes to an adult day program on Wednesdays through [program]. They have been fabulous. Absolutely fabulous. They’re very good. They phone to see if we need groceries or anything at all because they’re willing to do it. And then tomorrow they said they were going around to all the clients and delivering baking. Every day of the program we always hear from them. Over the phone. Yeah.” (Participant 111; Care Partner)*

#### Virtual Medicine: barriers and facilitators

Due to the scale of immediate shift to social and physical distancing, health care providers and patients had to quickly adapt to remote delivery of care in a virtual medicine format. Overall, many participants expressed that although they prefer face to face appointments with their care team, there were perceived advantages to receiving care in a remote format. These included not feeling as rushed, having options presented such as telephone vs zoom, and care partners expressed an ability to be more candid with the doctor if the person living with dementia was not participating in the call.

> *“And that was… we felt that that was good. My husband wasn’t part of the conversation because we were discussing some test results. We asked, I did ask my husband if he wanted to be part of the conversation or if he was comfortable that I would relay the information. And so, um, because he chose to not be part of the conversation, the doctor and I could be a little more candid.” (Participant 106; Care Partner)*
>
> *“But the conversation was very good because we both had time to ask questions, answer the questions, and I would say my doctor probably, hopefully, didn’t feel rushed. And I didn’t feel rushed. And I think that part was very much a bonus.” (Participant 106; Care Partner)*
>
> *“It’s going to be over Zoom. Oh, we had one yesterday with [nurse]. Did, a bunch of testing yesterday and that worked just fine. Actually I think it worked out great. He was able to do the drawings and then hold the paper up and show her- I think it worked fabulous.” (Participant 129; Care Partner)*

Conversely, care partners did also report that if the person living with dementia was in the room or wanted to be part of the call there was less opportunity for frank one on one conversations with the doctor and that this could be potentially distressing for the person living with dementia, depending on the nature of the discussion. Participants also expressed some concern with technological barriers of connecting via internet and while recognizing that body language and non-verbal communication was important to the appointment, support is necessary for people who are unfamiliar with the technology to be able to fully participate in this way.

> *“…for this next month, I’ve actually written a letter and I put it on a spreadsheet. I’m hoping I don’t have to go over it verbally with him [the doctor] and he can read the spreadsheet and then we can talk about it.” (Participant 143; Care Partner)*
>
> *“Like my mom had a follow up, uh, doctor’s appointment that was, at least they’re gonna do it on the phone as opposed to on a Zoom session. Because I hear this from my dad a lot, it’s like, you know what? Everybody wants to do all this stuff online and, I mean, some, some seniors capability of using computers is a little more limited.” (Participant 126; Care Partner)*

### Health Status Impact of Public Health Measures

#### Anxiety and Mental Health Concerns

Participants described feeling anxiety about the pandemic in general and about needing to leave the house, both personally and perceived anxiety on the part of the person living with dementia. This highlights the need to be cognizant of increased mental health needs for cognitive clinic patients and family members in times of social and physical distancing.

> *“Well, I do find that he is definitely more anxious and worried and fearful, but I have no idea if that’s related. I doubt that it is. I imagine that it’s more just a progression of the dementia.” (Participant 105; Care Partner)*
>
> *“Well now on the phone, he’s asking me the same questions and [Person living with dementia]’s right there with me because that’s what he wants. And so that was awful…because [Person living with dementia] gets very upset when we talk about stuff…” (Participant 143; Care Partner)*
>
> *“Yeah, and it’s so difficult… That taught me a lesson. So we went out. I couldn’t leave it, so I have to go pick it up today. And that’s frustrating because that gives you more anxiety of just leaving the house. I don’t have trouble leaving the house, but if [Person living with dementia] comes with me he has to wait in the car. And then, it was just really stupid of me. I said my brain’s going under too, a huge thing. Nothing’s in sync.” (Participant 111; Care Partner)*

#### Social distancing and cognitive decline

A number of participants also expressed concern that the person living with dementia was experiencing more cognitive decline since the start of the public health measures during the COVID-19 pandemic. These concerns were focused around the decrease in social interaction impacting the progress and severity of cognitive decline, which was perceived as progressing more rapidly the previous two months.

> *“I mean, my mom didn’t have great recollection of going to [day program], I recall. Like when I’d visit her she’d vaguely remember doing stuff. I don’t know if it provided a different level of stimulus because I seem to have noticed a little bit more of a… kind of slow decline or decay… Where maybe it’s not getting the same activity now. Um, so noticing a slight decline in her cognition… I mean if the decline is a slope, the slope’s a little bit steeper in the last couple months than it was…” (Participant 126; Care Partner)*
>
> *“Her [centre] program was something that I think she really thrived when she was going there, especially I noticed, you know, mood improvement, sleep improvement, all of those types of things. The more activity that my mom is doing… the better off. She was swimming every Saturday before this. Now, I feel like with changes that have kind of happened, those types of things, I think, will be hard to get back into that type of an exercise environment, which sucks because that was something that if you keep in the routine and don’t change, then I think it could have gone on for a lot longer. I think with the decline that’s kind of happened over the past five weeks, those types of things will have to be removed, unfortunately. I found that, in the past five weeks, her telephone skills have, like, literally disappeared. That is non-existent. She was able to pick up the phone and, you know, say ‘Hello’, all that kind of stuff, non-existent now.” (Participant 124; Care Partner)*

#### Care partner burnout

Many care partners expressed feeling as though they could not cope on their own, with limited access to both formal and informal supports. The changes to the health systems and services provided described in the section above have effects on the amount of burnout expressed by family members and care partners. Participants also worried about returning to work if the services that enabled them to do so pre-pandemic were not reinstated.

> *“Well, I don’t know how it works with social distancing, but I am, we have had quite a decline. I’m not saying it’s from this social distancing, but there’s been a marked decline… I’m on the verge of asking for someone coming in despite the fact that we’re in social, you know, it’s been a lot. It would be helpful if they could, I guess ease up and have a visitor come around once a week or something and have some time where you just had an hour when you weren’t on… on call 24/7.” (Participant 143; Care Partner)*
>
> *“My father’s providing primary care duties and they’re in their 80s type thing. One of the key things that’s changed is that my mom used to go to day program twice a week that gave my dad some respite and that kind of obviously stopped. So that’s put more stress on him because he’s not getting that relief type thing. And my dad doesn’t have a big social network to begin with and, uh, that’s been restricted by the, the kind of public health measures and, and the social distancing. And, uh, that’s made things more difficult. I mean, he’s doing the primary caregiving. Um, he’s doing all the, the kind of, there’s, there’s no rest but he’s doing all the work at home in terms of meal preparation, things like that… that’s put a lot of stress on my dad. He’s not been given a lot of great resources.” (Participant 126; Care Partner)*
>
> *“I lost all my support. All of it, except for a few hours. So I was just left to just manage this on my own. Yeah… you know, it, going from, you know, 24 hours of day program and 35 hours of home care to nothing. They stripped all my support away… Yeah. Like, I am scheduled to start going back to work again and no one, no one can tell me what I’m supposed to do with my spouse, because the day programs are no longer active. So for 24 hours a week now I don’t know what I’m supposed to do.” (Participant 141; Care Partner)*

## Discussion

This study provides early evidence of the impact of COVID-19 on persons living with dementia and their care partners. These interviews identified multiple adverse consequences of the public health measures implemented to contain spread of SARS-CoV-2, including loss of informal caregiving, lack of access to health care, and social isolation, contributing to burn out symptoms among care partners. This validates the concerns expressed by multiple advocacy organizations (such as the Alzheimer Society of Canada, Alzheimer’s Association (USA), and Alzheimer’s Disease International), which although well founded have nonetheless been based on anecdotes or speculation without data collected directly from persons living with dementia. Our data were collected during a period of “lockdown” when all non-essential businesses were closed, health clinic access was mandated to be online or telephone except for emergencies, and most in-person home care services were no longer being provided. Therefore, it provides a snapshot of the adverse effects of the aggressive public health efforts that were needed to “flatten the curve” to prevent health system collapse. These drastic public health measures, while being lightened in many jurisdictions, may yet be required in other countries at an earlier stage of the pandemic or in case of a second wave of viral infections in the Northern hemisphere in the fall.

So far, there are few data on the impact of COVID-19 on dementia care even though it is well understood that older people with medical comorbidities are at much higher risk of fatal infection. In a letter in the Lancet, physicians from Wuhan province cited multiple barriers to good dementia care including difficulty adhering to social distancing and good hygiene, loss of services, and difficulty accessing telemedicine (Wang et al., 2020). However, no perspective from persons living with dementia or quantitative information was included. A hospital-based cohort from Italy noted that patients with COVID-19 infection with a history of dementia were more likely to die, and often presented with hypoactive delirium (Bianchetti et al., 2020). A quantitative survey study of community dwelling persons with mild cognitive impairment or mild dementia in Spain reported overall good mental health (Goodman et al., 2020). However, that study used a quantitative survey without open-ended questions, did not include care partners, and was limited to persons with mild dementia. In contrast, this study elicited many serious concerns particularly among care partners of persons with more moderate to severe dementia.

We identified multiple impacts on well-being of people living with dementia and their family members and/or care partners. Though most participants understood the need for social distancing, they found it difficult both physically and emotionally. Meaningful activity and social engagement are vital to the well-being and functioning of families of persons living with dementia (Phinney, 2006; Phinney et al., 2007; Roach & Drummond, 2014; Roach et al., 2016). Care partners expressed concerns that cognitive decline accelerated during social distancing. This is consistent with literature finding that loneliness can be detrimental to well-being and quality of life (Moyle et al., 2011) and increase the risk of mortality and morbidity (Luo et al., 2012; Tilvis et al., 2012). Social interaction can be beneficial for cognitive functioning (Vernooij-Dassen & Jeon, 2016) and so it is plausible that social isolation could provoke accelerated cognitive decline and or an exacerbation of neuropsychiatric symptoms of dementia.

The adverse impact of the COVID-19 pandemic on mental health is increasingly recognized (Armitage & Nellums, 2020; Holmes et al., 2020). These mental health impacts may be compounded when care partners no longer have access to formal and informal supports that provide respite, help with hygiene, medication, day to day activities of living, and essential tasks such as shopping for groceries or collecting medications. Moreover, the challenges of assisting a person living with dementia to navigate changing public spaces where physical distancing, wearing of masks, and controlled movements are necessary leads to additional stress for the care partner and greater risk for the person living with dementia.

Participants were understanding of these measures but at the same time expressed concern for their own ability to continue to cope on a long-term basis, and the possibility of burnout without these supports. This echoes general concern of burnout by families living with dementia in times before the COVID-19 pandemic (Brodaty & Donkin, 2009; Sörenson & Conwell, 2011; Takai et al., 2009), but with these caring demands now amplified by isolation and anxiety. With the accompanying sudden shift to virtual care by almost all health care teams there was additional anxiety experienced with regards to the availability of care and how to access it.

Many of the stresses and anxieties identified in this study could potentially be mitigated through health system innovations. Remote support for social interaction for people living with dementia and care partners should be explored. There is evidence that tablet based interventions can benefit cognition and self-perceived quality of life for people living with dementia (Hung et al., 2020; Kong, 2020) and ongoing work is being made with integrating social robots into the care of people with dementia in the community (Hung et al., 2019; Korchut, 2017). Rapid implementation of these innovative technological solutions may provide one strategy to increase social interaction and improve well-being in times of pandemic public health restrictions. Additional evidence is needed to demonstrate what kind of interaction may be valuable for these approaches, and what types support may be most facilitative of a protective effect for cognition (Kim et al., 2015).

Other possible remote support for people living with dementia and families include virtual social networking, including virtual dementia cafes or day programming moving to online interaction (Hung & Mann, 2020). Where this is not possible, telephone check-ins may also prove to be a valuable source of support for the mental health and needs assessment for families. The more problematic question remains focused on remote approaches to care partner respite, which is difficult to navigate during times of public health restrictions. Possible solutions for home care or social care workers may include scheduled home care visits incorporating walks where the formal care provider and person living with dementia maintain an appropriate social distance; meal or medication collection and delivery at a safe distance or with the appropriate personal protective equipment, and flexibility on maximum limits for dispensing medications to the reduce the number of needed trips to the pharmacy.

Delivering high quality telemedicine will be essential to ensuring good health during a time of social distancing. It was evident in the data that if patients and care partners are expected to connect with internet technology, clear guidance should be provided to explain how to set up and use technology that may be unfamiliar to many. The ability for a care partner to provide confidential information to a clinical care team in advance of a scheduled virtual appointment was also expressed, particularly when the care partner wished to notify the medical team of symptoms that are potentially distressing or humiliating to the patient, or could lead to conflict. To address this need, our clinic is implementing secure web-based surveys that will allow the care partner to confidentially report neurobehavioural symptoms, function, and caregiver burden, using validated scales, with separate telemedicine appointments with the care partner as necessary. This work leverages data systems employed to capture patient data in the PROMPT registry, demonstrating another use of clinical patient registries for meaningful patient engagement and service improvement. Previous work has shown that neurological and psychiatric patient populations do not see registry participation as burdensome, but as an altruistic way to contribute to research that may or may not also provide personal benefit (Lee et al., 2019). Enhancing communication and preparation for clinic appointments has been shown to improve health outcomes in non-pandemic times (Entwisthle & Watt, 2006; Street et al., 2009) and we can reasonably extrapolate that this would be true during times of social and physical distancing, as experienced during the COVID-19 pandemic.

## Strengths and Limitations

Strengths of the study include that it was embedded in a prospective clinic registry which enabled us to quickly contact people living with dementia and their care partners during the COVID-19 pandemic, and link their data to physician collected information on cognitive assessment and disease diagnosis. The main limitation is that the study was conducted in a single setting, a specialty outpatient dementia clinic, in an urban location with universal healthcare. It may not be wholly applicable to persons receiving care in family practices or with limited access to care. In future work we will also work to include more people living with dementia in the interviews, and perhaps the necessity of remote interviewing in this study presented a barrier to communication. However, the themes that emerged from our work accord with the concerns expressed by multiple advocacy groups, expert opinion (Brown et al., 2020), and the early observations of dementia specialists in Wuhan province (Wang et al., 2020); therefore, we expect that most of the themes are universal and not idiosyncratic to our practice setting. By design, we did not include persons living with dementia who are residing in care homes or who are hospitalized in acute care; additional research is needed on these important populations. This work is limited to the impacts of the initial public health response to the COVID-19 pandemic, and it will be useful to continue additional qualitative work to better understand the impact on different groups over time as the public health response evolves and society adapts to the changes.

## Clinical Implications

This in-depth understanding of the lived impact of decreased social engagement and healthcare access on personal well-being can inform future health and social policy and health service provision. This knowledge may be useful for future pandemics (including the possibility of a resurgence of COVID-19 in the fall); extreme weather phenomenon; outbreaks of other disease or isolation in supportive living or long term care facilities; or other emergencies where social distancing/isolation/virtual care may be required. The reported increase in neuropsychiatric symptoms is also important for treatment planning and support in times of social distancing. Improved and innovate approaches to remote care and virtual medicine, including flexible mental and social supports for families, and the ability to provide information to clinical care teams prior to remote medical appointments has the potential to facilitate high-quality patient care that meets the needs of families and patients.

## Data Availability

Qualitative interview data is available as provided in manuscript

## Acknowledgements

The PROMPT registry is funded by the Katthy Taylor Chair in Vascular Dementia of the University of Calgary, with support from the Brain and Mental Health Research Clinics Initiative funded by the Hotchkiss Brain Institute, Department of Clinical Neurosciences, and Department of Psychiatry at the University of Calgary. The PROMPT Collaborators are: Drs. Eric Smith, Zahinoor Ismail, Dawn Pearson, Aaron Mackie, Alicja Cieslak, Brienne McLane, Bijoy Menon, Dallas Seitz, David Patry, Phil Barber, Robert Granger, David Hogan, and Collen Maxwell.

## References

Alzheimer’s Disease International. (2020). ADI offers advice and support during COVID-19. https://www.alz.co.uk/news/adi-offers-advice-and-support-during-covid-19 (accessed May 29, 2020).

Armitage, R. & Nellums, L.B. (2020). COVID-19 and the consequences of isolating the elderly. The Lancet Public Health, 5: e256.

Bianchetti, A., Rozzini, R., Guerini, F., Boffelli, S., Ranieri, P., Minelli, G., Bianchetti, L., & Trabucchi, M. (2020). Clinical Presentation of COVID19 in Dementia Patients. The Journal of Nutrition, Health & Aging (doi: 10.1007/s12603-020-1389-1).

Braun, V., Clarke, V., & Weate, P. (2016). Using thematic analysis in sport and exercise research. In: B. Smith and A. C. Sparkes, eds., International handbook on qualitative research in sport and exercise. London: Routledge, pp. 191–218.

Brodaty, H. & Donkin, M. (2009). Family caregivers of people with dementia. Dialogues in Clinical Neuroscience, 11(2): 217–228.

Brown, E.E., Kumar, S., Rajji, T.K., Pollock, B.G., & Mulsant, B.H. (2020) Anticipating and Mitigating the Impact of the COVID-19 Pandemic on Alzheimer’s Disease and Related Dementias. American Journal of Geriatric Psychiatry (doi: 10.1016/j.jagp.2020.04.010).

Clarke, V., Braun, V., Terry, G., & Hayfield, N. (2019). Thematic analysis. In: P. Liamputtong, ed., Handbook of research methods in health and social sciences. Singapore: Springer, pp. 843–860.

Dewing, J. (2007). Participatory research: A method for process consent with persons who have dementia. Dementia: the international journal of social research and practice, 6, pp.11–25.

Entwistle, V.A. & Watt, I.S. (2006). Patient involvement in treatment decision-making: The case for a broader conceptual framework. Patient Education and Counseling, 63: 268–278.

Feeney, B.C. & Collins, N.L. (2015). A New Look at Social Support: A Theoretical Perspective on Thriving Through Relationships. Personality and Social Psychology Review, 19(2), pp.113–147.

Folstein, M. F., Folstein, S. E., & McHugh, P. R. (1975). Mini-mental state: A practical guide for grading the cognitive state of patients for the clinician. Journal of Psychiatric Research, 12: 189–198.

Goodman-Casanova, J.M., Dura-Perez, E., Guzman-Parra, J., Cuesta-Vargas, A., & Mayoral-Cleries, F. (2020). Telehealth Home Support During COVID-19 Confinement for Community-Dwelling Older Adults With Mild Cognitive Impairment or Mild Dementia: Survey Study. Journal of Medical Internet Research, 22: e19434. (doi: 10.2196/19434).

Holmes, E.A., O’Connor, R.C., Perry, V.H., Tracey, I., Wessely, S., Arseneault, L., Ballard, C., Christensen, H., Silver, R.C., Everall, I., Ford, T., John, A., Kabir, T., King, K., Madan, I., Michie, S., Przybylski, A.K., Safran, R., Sweeney, A., Worthman, C.M., Yardley, L., Cowan, K., Cope, C., Hotopf, M., & Bullmore, E. (2020). Lancet Psychiatry, 7: 547–560.

Hung, L., Chow, B., Shadarevian, J., O’Neill, R., Berndt, A., Wallsworth, C., Horne, N., Gergorio, M., Mann, J., Son, C., & Chaudhury, H. (2020). Using touchscreen tablets to support social connections and reduce responsive behaviours among people with dementia in care settings: A scoping review. Dementia: the international journal of social research and practice (doi: http://dx.doi.org/10.1177/1471301220922745).

Hung, L. & Mann, J. (2020). Virtual special issue – Using touchscreen tablets for virtual connection. Dementia: the international journal of social research and practice (doi: http://dx.doi.org/10.1177/1471301220924578).

Hung, L., Berndt, A., Wallsworth, C., Horne, N., Gregorio, M., Mann, J., Liu, C., Woldum, E., Au-Yeung, A., & Chaudhury, H. (2019). Involving patients and families in a social robot study. Patient Experience Journal, 6(2): article 12 (doi: 10.35680/2372-0247.1362).

Kim, G.H., Jeon, S., Im, K., Kwon, H., Lee, B.H., Kim, G.Y., Jeong, H., Han, N.E., Seo, S.W., Cho, H., Noh, Y., Park, S.E., Hojeong, K., Hwang, J.W., Yoon, C.W., Kim, H.J., Byoung, S.Y., Chin, J.H., Kim, J-H., Suh, M.K., Lee, J.M., Kim, S.T., Choi, M-T., Kim, M.S., Heilman, K.M., Jeong, J.H., & Na, D.L. (2015). Structural Brain Changes after Traditional and Robot-Assisted Multi-Domain Cognitive Training in Community-Dwelling Healthy Elderly. PLoS ONE, 10(4): e0123251.

Kitwood, T. (1997). Dementia Reconsidered: the person comes first. Buckingham: Open University Press.

Kong, A.P. (2020). The use of free non-dementia-specific Apps on iPad to conduct group communication exercises for individuals with Alzheimer’s disease (Innovative Practice). Dementia: the international journal of social research and practice, 19(4): 1252–1264.

Korchut, A., Szklener, S., Abdelnour, C., Tantinya, N., Hernández-Farigola, J., Ribes, J.C., Skrobas, U., Grabowska-Aleksandrowicz, K., Szcześniak-Stańczyk, D., & Rejdak, K. (2017). Challenges for Service Robots—Requirements of Elderly Adults with Cognitive Impairments. Frontiers in Neurology, 8: article 228 (doi: 10.3389/fneur.2017.00228)

Lee, J.Y.Y., Crooks, R.E., Pham, T., Korngut, L., Patten, S., Jetté, N., Smith, E.E., & Roach, P. (2019). “If it helps someone, then I want to do it”: Perspectives of persons living with dementia on research registry participation. Dementia: the international journal of social research and practice (doi: http://dx.doi.org/10.1177/1471301219827709).

Lincoln, Y. S. & Guba, E. G. (1985). Naturalistic Inquiry. Newbury Park: Sage.

Luo, Y., Hawkley L.C., Waite L.J., & Cacioppo J.T. (2012). Loneliness, health, and mortality in old age: A national longitudinal study. Social Science & Medicine, 74, pp.907–14.

Moyle, M., Kellett, U., Ballantyne, A., & Gracia, N. (2011). Dementia and loneliness: an Australian perspective. Journal of Clinical Nursing, 20: 1445–1453.

Phinney, A. (2006). Family strategies for supporting involvement in meaningful activity by persons with dementia. Journal of Family Nursing, 12, 80–101.

Phinney, A., Chaudhury, H., & O’Connor, D. (2007). Doing as much as I can do: The meaning of activity for people with dementia. Aging & Mental Health, 11, pp.384–393.

QSR International Pty Ltd. (2018). NVivo qualitative data analysis software, version 12.

Qualtrics. (2019). Qualtrics, Provo, UT, USA. https://www.qualtrics.com

Roach, P., Drummond, N., & Keady, J. (2016). ‘Nobody would say that it is Alzheimer’s or dementia at this age’: Family adjustment following a diagnosis of early-onset dementia. Journal of Aging Studies, 36, pp.26–32.

Roach, P. & Drummond, N. (2014). ‘It’s nice to have something to do’: early-onset dementia and maintaining purposeful activity. Journal of Psychiatric and Mental Health Nursing, 21, pp. 889–895.

Sheikh, F., Ismail, Z., Mortby, M.E., Barber, P., Cieslak, A., Fischer, K., Granger, R., Hogan, D.B., Mackie, A., Maxwell, C.J., Menon, B., Mueller, P., Patry, D., Pearson, D., Quickfall, J., Sajobi, T., Tse, E., Wang, M., & Smith, E.E. for the PROMPT registry investigators. (2018). Prevalence of mild behavioral impairment in mild cognitive impairment and subjective cognitive decline, and its association with caregiver burden. International Psychogeriatrics, 30: 233–244. (doi: 10.1017/S104161021700151X).

Sörenson, S. & Conwell, Y. (2011). Issues in dementia caregiving: Effects on mental and physical health, intervention strategies, and research needs. American Journal of Geriatric Psychiatry, 19(6): 491–496.

Street Jr., R.L., Makoul, G., Arora, N.K., & Epstein, R.M. (2009). How does communication heal? Pathways linking clinician patient communication to health outcomes. Patient Education and Counseling, 74: 295–301.

Takai, M., Takahashi, M., Iwamitsu, Y., Ando, N., Okazaki, S., Nakajima, K., Oishi, S., & Miyaoka, H. (2009). The experience of burnout among home caregivers of patients with dementia: Relations to depression and quality of life. Archives of Gerontology and Geriatrics, 49(1): e1-e5.

Tilvis, R.S., Routasalo, P., Karppinen, H., Strandberg, T.E., Kautiainen, H., & Pitkala, K.H. (2012). Social isolation, social activity and loneliness as survival indicators in old age; a nationwide survey with a 7-year follow up. European Geriatric Medicine, 3, pp.18–22.

Vernooij-Dassen, M. & Jeon, Y-H. (2016). Social health and dementia: the power of human capabilities. International Psychogeriatrics, 28(5), pp.701–03.

Wang, H., Li, T., Barbarino, P., Gauthier, S., Brodaty, H., Molinuevo, J.L., Xie, H., Sun, Y., Yu, E., Tang, Y., Weidner, W., & Yu, X. (2020). Dementia care during COVID-19. Lancet, 395: 1190–1191. (PMID 32240625).

Ward, R., Clark, A., Campbell, S., Graham, B., Kullberg, A., Manji, K., Rummery, K., & Keady, J. (2018). The lived neighbourhood: understanding how people with dementia engage with their local environment. International Psychogeriatrics, 30(6), pp.867–80.

